# Encouraging healthy weight for adults with intellectual disabilities: supporting the role of carers

**DOI:** 10.1101/2025.10.22.25338600

**Authors:** Karen Coulman, Justine Womack, Carly Atkinson, Briony Caffrey, Alison Tavaré, Nicola Powell, Roz Erskine-Gray, Rachel Roberts, Nick Pugh, Amanda Owen-Smith

**Author notes:** Corresponding author: Karen Coulman, Population Health Sciences, Bristol Medical School, University of Bristol, 1-5 Whiteladies Road, Bristol, BS8 1NU, UK.

## Abstract

Adults with intellectual (learning) disabilities die on average 20 years earlier than the UK general public, with heart disease a leading cause of avoidable death. Obesity, a key risk factor, is more prevalent in this group. Carers (paid and unpaid) play a vital role in supporting healthy behaviours and healthy weight in adults with intellectual disabilities but often lack appropriate training.

This study developed national e-learning for carers based on qualitative research methods. This included 1) content analysis of professional resources to promote healthy weight in people with intellectual disabilities, 2) semi-structured interviews with health and social care managers, clinical leads and commissioners to identify carer barriers and areas of training need, 3) semi-structured interviews and focus groups with paid and unpaid carers to feedback on and refine training content. Thematic analysis of interview and focus group transcripts was undertaken to identify key themes Twenty-one professional resources and 25 participants (across 19 interviews and two focus groups) informed the training. Two overarching concepts explained themes arising from the data - the central yet under-supported role of carers in promoting healthy weight, and the structural and cultural barriers to healthy weight in people with intellectual disabilities. Identified carer training needs included: the health inequalities faced by people with intellectual disabilities, causes and health impacts of obesity, healthy eating and physical activity recommendations, and how to apply these topics when working with an adult with an intellectual disability. The research team worked with an intellectual disability charity ‘experts by experience’ group to develop lived experience videos of individuals’ journeys towards healthier weight included within the e-learning.

Carers face many barriers in supporting healthy behaviours in people with intellectual disabilities, including limited knowledge of obesity, nutrition and physical activity. National e-learning was developed from this research, available at: https://www.e-lfh.org.uk/programmes/supporting-healthy-weight-in-an-adult-with-a-learning-disability/.

## Background

People with intellectual disabilities (also referred to as ‘learning disabilities’) face significant health inequalities and have a life expectancy 20 years shorter than the United Kingdom (UK) general population (1). The latest LeDeR (Learning from lives and deaths – People with a learning disability and autistic people) report found that 39% of deaths were ‘avoidable’, compared with 22% in the general population, with cardiovascular disease a leading cause (1). Intersecting inequalities worsen outcomes, for example people with intellectual disabilities from ethnic minority backgrounds have an average life expectancy of just 34 years, half that of their White counterparts (2).

Obesity is more prevalent in people with intellectual disabilities, and is a key risk factor for cardiovascular disease, amongst other health conditions (3–6). Contributing factors include poor diet and sedentary lifestyle in part related to being disproportionately affected by poverty, deprivation, social isolation, and the presence of conditions or medications which pre-dispose to weight gain (3, 7, 8).

Addressing obesity is a key priority for reducing avoidable deaths in people with intellectual disabilities. A recent Nuffield Trust report identified obesity as one of five focus areas to close the gap of health inequalities with the general population (9). Recommendations include improving access to preventative health services, and better coordination of care between health and social care settings including consistent training of health and care professionals.

Previous literature highlights the vital role of carers, both paid and unpaid, in supporting healthy behaviour change in people with intellectual disabilities (10–14). A 2024 realist review by Westrop *et al* identified carer involvement as essential to lifestyle modification in people with intellectual disabilities, with carers’ life and work pressures important contexts affecting their ability to provide support (14). A systematic review using a socio-ecological model found limited carer training and health literacy to be major barriers to people with intellectual disabilities achieving a healthy eating pattern and weight loss (12). Westrop *et al* also called for training interventions targeting carers “*to help mediate these contexts and have positive impact on lifestyle change*” (14). In supporting a person with an intellectual disability to engage with a weight management programme, carers have stressed the importance of accessible quick-to-read materials that fit within the time pressures of their role (13).

This research aimed to 1) investigate the key training needs for carers in supporting healthy weight in adults with intellectual disabilities, and 2) develop free nationally available e-learning for carers on supporting healthy weight in adults with intellectual disabilities.

## Materials and Methods

Qualitative research methods were used to address the study aims across three main stages (**Figure 1**). The project was informed by and used techniques from Grounded Theory, focusing on an inductive emergence of themes from the data (15). The research team consisted of academics (KC and AOS), multi-disciplinary health professionals (KC, CA, BC, R-EG, NPo, AT, JW), regional policy leads (JW, NP), a parent carer of an adult with an intellectual disability (R-EG), and professionals from an intellectual disability charity (RR, NPu). This study is reported according to the Consolidated criteria for reporting qualitative research (COREQ) **(Appendix 1)** (16).

**Figure 1:**
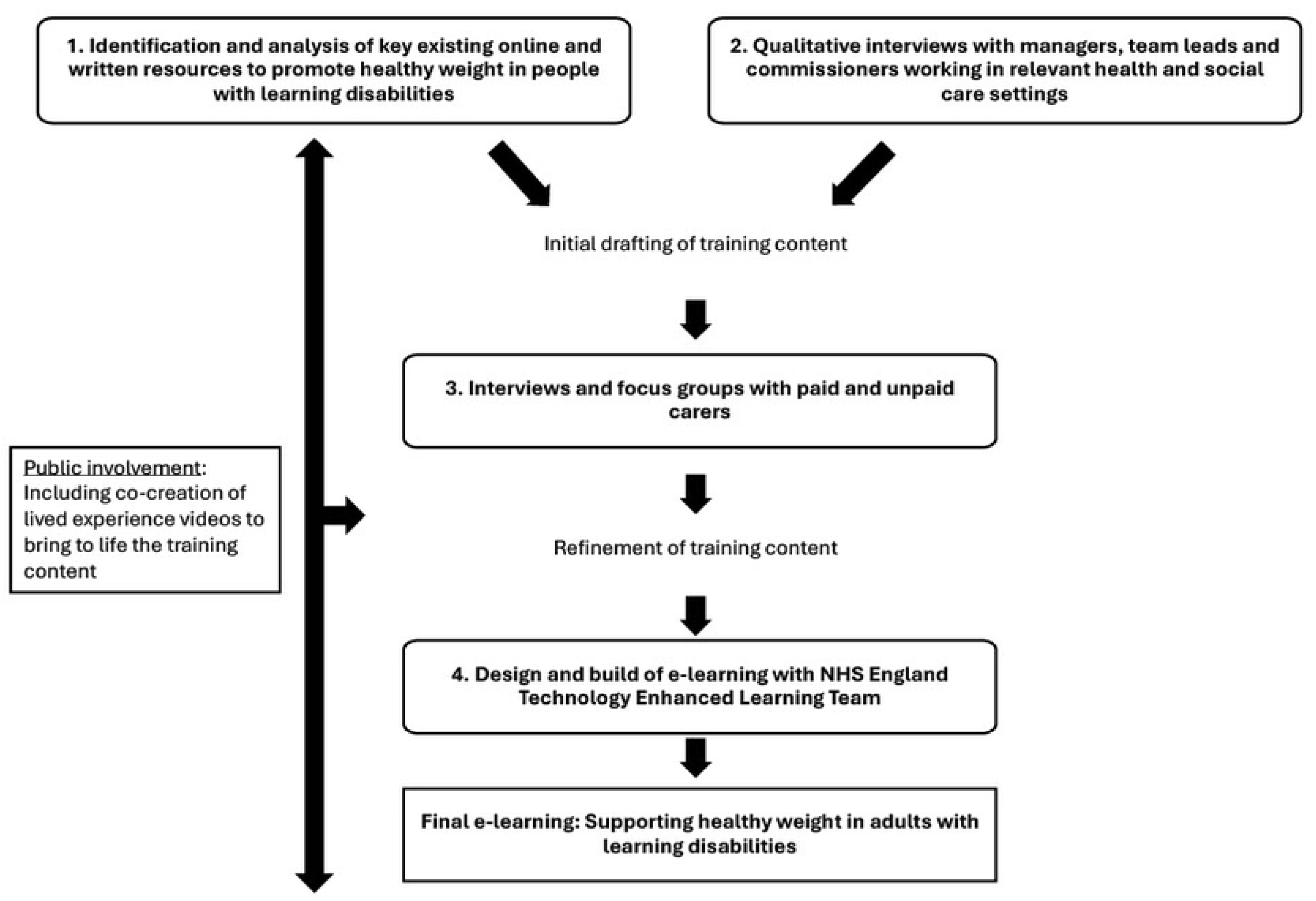
Overview of research methods to develop national e-learning for carers on supporting healthy weight in an adult with a learning disability

### Ethics statement

This research received ethical approval from the Faculty of Health Sciences Research Ethics Committee at the University of Bristol (Ref. 15326). All study participants provided written informed consent prior to their inclusion in the study.

### 1. Identification and analysis of existing professional resources to promote healthy weight in people with intellectual disabilities

The aim of stage 1 was to identify existing written and online resources used by UK professionals to promote healthy weight in people with intellectual disabilities and to identify the topics covered by these resources. Resources were identified in three ways:

1. Targeted internet searches of relevant professional organisations’ online resources (e.g. Mencap).
2. Requests for relevant resources were sent through the South West of England healthy weight and learning disability community of practice email list and at community of practice meetings.
3. A review of resources available or signposted from key professional UK organisations involved in supporting people with intellectual disabilities and their carers (including the British Dietetic Association, the Learning Disability Senate and the Caroline Walker Trust).

#### Data collection and analysis

All identified resources were catalogued in an Excel spreadsheet. A rapid thematic analysis of each resource was undertaken (17). Initially each resource was categorised according to its main focus (e.g. Healthy eating, physical activity). Each resource was then scrutinised by the lead researcher (KC a qualitative researcher and registered dietitian) to identify the topics covered in each resource. Lists of topics from each resource were then combined to create one comprehensive list of topics. These were grouped into themes based on similarity of topics, with any duplicates removed to create a final list of categorised topics.

### 2. Qualitative interviews with health and social care managers, clinical leads, and commissioners

The aim of stage 2 was to investigate the views of managers and commissioners responsible for delivering services for people with intellectual disabilities about areas of training need for supporting people with intellectual disabilities with healthy weight.

#### Sampling

Sampling was carried out purposively using a key informant approach starting with key members of the South West healthy weight learning disability community of practice, and relevant professional contacts of the research team (18, 19). We aimed to include a range of informants from different professional backgrounds and sectors working in a team lead, management or commissioning role for services for people with intellectual disabilities. Potential participants were contacted by their professional contact via email enclosing an information sheet about the research. They were asked to respond directly to the main researcher (KC) if interested. KC responded to interested participants by phone or email to discuss and answer questions about the research and arrange a time and location for the interview. Sampling and recruitment, undertaken in tandem with data collection and analysis, continued until the research team agreed that sufficient ‘information power’ had been obtained relative to the aims of the study (20).

#### Data collection and analysis

Semi-structured individual interviews were used to elicit participants’ accounts of planning and managing healthy weight interventions for people with intellectual disabilities. The full interview topic guide is available in **Appendix 2**. Participants were offered the option of conducting the interview in-person, online (via Microsoft Teams) or via telephone. KC, a female experienced qualitative researcher and registered dietitian specialising in obesity conducted all interviews and focus groups across stages 2 & 3.

All interviews were audio-recorded and fully transcribed. Key notes relating to topics to follow-up on in subsequent interviews were written up by the researcher after interviews. A thematic analysis of interview transcripts was undertaken, using techniques of constant comparison to code data and identify emerging themes (15). Coding and management of the data was undertaken by KC and facilitated using NVivo 12 software (21). Themes emerging from the data were written up as detailed descriptive accounts, and matrices drawn up to facilitate the inter-relation of themes and make comparisons across interviews (22). Codes and emerging themes were presented and discussed with the research team at monthly team meetings. Data analysis ran in parallel with sampling and data collection to allow emerging themes to be followed up and synthesised, triangulating between the views of different participants throughout (15). Negative cases, where informants held divergent views or experiences did not follow the standard course of events, were re-analysed to gain further insights.

#### Initial drafting of training content

Early in the study, meetings were held between KC and the National Health Service England (NHSE) Technology Enhance Learning (TEL) team to discuss and provide advice on the process of translating research findings into accessible training content. An initial draft of the training structure and content informed by research stages 1&2 was developed by KC with input from the study team at steering group meetings. Content was presented on PowerPoint slides which were used in stage 3 of the research.

### 3. Qualitative interviews and focus groups with paid and unpaid carers

The aim of stage 3 was to investigate the acceptability of the draft training content from the perspective of paid and unpaid carers.

#### Sampling

Professional (paid) carers/support workers and family (unpaid) carers were identified via key informants interviewed in stage 2, and through professional contacts of the research team. Key informants were asked to pass on information about the research to potential participants. Interested individuals contacted KC directly, and arrangements made as described for stage 2.

#### Data collection and analysis

Semi-structured individual interviews and focus groups were used to elicit carers’ views and experiences of supporting healthy weight in people with intellectual disabilities, as well as their reflections on the draft training content developed. The topic guide is available in **Appendix 3**. Individuals took part in either an individual interview <u>or</u> focus group depending on preference. As per stage 2, participants were given the option of conducting the interview/focus group in-person or online (or via telephone for individual interviews). Recording, transcription and thematic analysis of interview and focus group transcripts was undertaken as described for stage 2.

### 4. Refinement of training content and design and build of e-learning

Training content was refined by KC based on findings from the carer interviews and focus groups, and with input from the study team. The refined training content was provided to the NHSE TEL team on PowerPoint slides. From the training content, the TEL team designed a content plan on PowerPoint slides which members of the research team fed back on. Once the content plan was agreed, a test version of the session was developed in the interactive platform which the research team tested and provided final feedback on.

### Public involvement

A family carer of an adult with a learning disability (RE-G) was part of the research team. She contributed to research team meetings, helped with the recruitment of carers to the research, and inputted into the development of the training content.

The research team also collaborated with the <u>Brandon Adventurers</u>, a panel of experts by experience from a learning disability charity that provides care and support for people with learning disabilities. The research was a topic of interest to the Brandon Adventurers who wished to contribute their stories to enhance the training. This was facilitated through RR and NP. We worked with the Adventurers to co-create lived experience videos to bring to life the training content included in the e-learning from the perspective of individuals with intellectual disabilities. These three two-minute videos feature the stories of four individuals with intellectual disabilities and their journeys with behaviour change and weight loss. KC worked with RR to develop a plan for the videos, based on the stories the four individuals wished to share. RR and NPu worked with the Adventurers to create the videos. Filming and editing were undertaken by NPu.

## Results

### Content analysis of resources

Twenty-one resources were identified and categorised according to their key focus as ‘Relevant national e-learning packages’ (n=4), ‘Other resources focusing on healthy eating guidance and/or physical activity’ (n=12), and ‘Meal planning and recipe resources’ (n=5) (**Appendix 4**).

Topics identified from the resources were categorised into five topic themes with 126 sub-themes: ‘Dietary/healthy eating information’ (48 sub-themes), ‘Physical activity information’ (13 sub-themes), ‘Supporting behaviour change’ (15 sub-themes), ‘Obesity/weight specific information’ (34 sub-themes) and ‘Capacity to make decisions/supporting independence’ (16 sub-themes). The full list of themes and sub-themes is available in **Appendix 5**.

### Qualitative interviews and focus groups

#### Participants

Twenty-five individuals took part in the research across 19 individual interviews and two focus groups (n=6). Participants included social care professionals (‘paid carers’, n=15, 60%), healthcare professionals (n=5, 20%), family carers (n=3, 12%), a public health professional (n=1, 4%), and a social enterprise professional (n=1, 4%). Participants came predominantly from the South West of England (n=22, 88%), with one from the North East of England (4%), and two holding national roles (8%). Participants were based in both urban and rural areas. Seventeen interviews and both focus groups were undertaken via Microsoft Teams, and two interviews via telephone. Interviews lasted between 41 and 61 minutes. Focus groups lasted approximately 90 minutes. All interviews and focus groups were conducted between December 2023 and September 2024.

An additional 18 individuals were approached about the research but did not take part; 12 initially expressed interest but did not get back to the researcher, and six did not get in touch with the researcher.

#### Thematic findings

Results are presented under five key themes identified from interviews and focus group discussions (**Figure 2**): 1. Challenges for healthy weight in adults with intellectual disabilities, 2. Accessing and engaging with ‘mainstream’ weight management services, 3. The role of the carer in supporting healthy weight, 4. Challenges for carers in supporting healthy weight, and 5. Carer training needs. Across these five themes, two overarching concepts were identified: 1. Carers as central but under supported agents in health promotion, and 2. Structural and cultural barriers to equitable weight management. The final NVivo codebook used in the analysis is available in **Appendix 6**.

**Figure 2:**
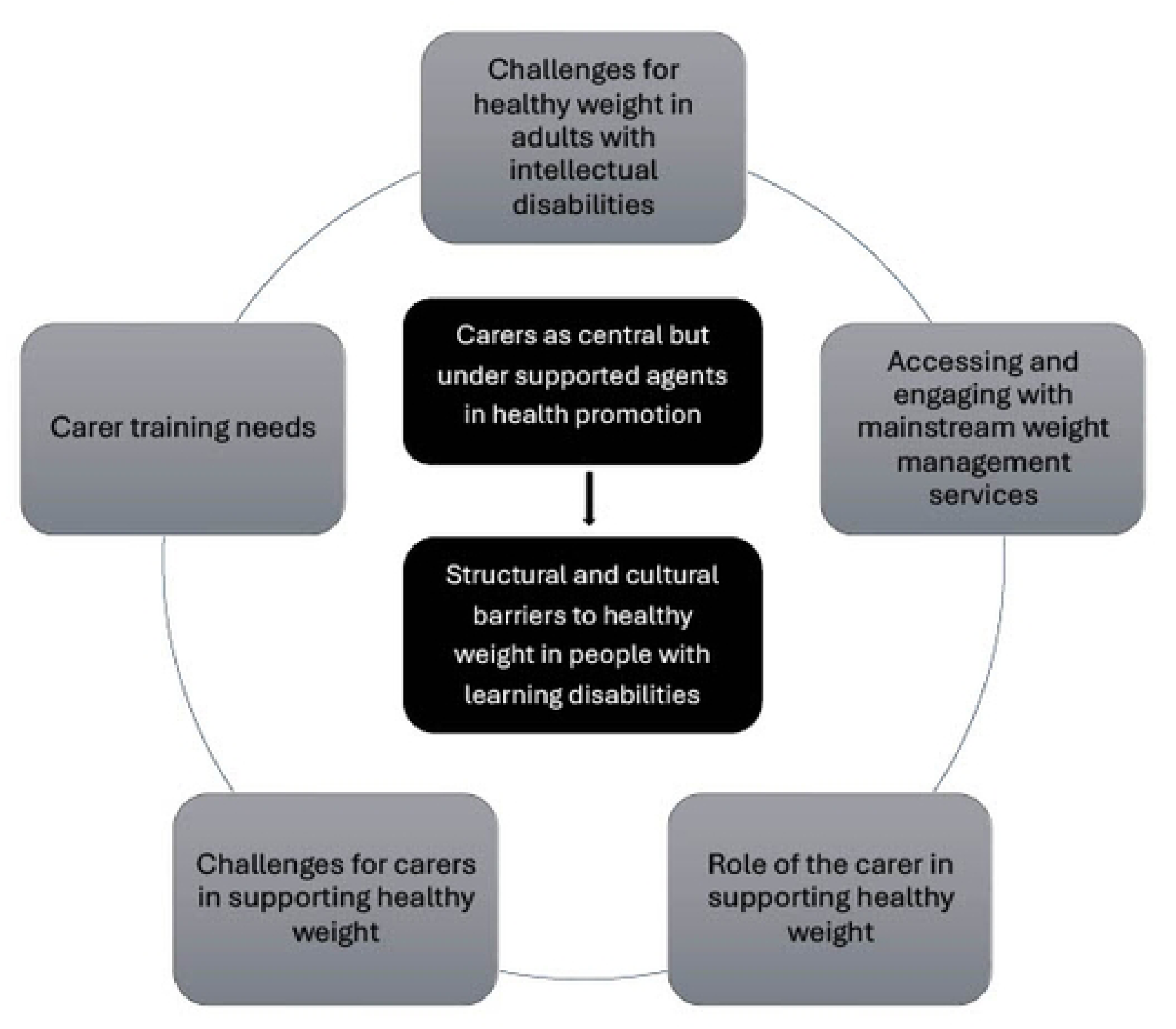
Themes and concepts relating to carers and supporting healthy weight in people with intellectual disabilities

##### Challenges for healthy weight in adults with intellectual disabilities

Participants highlighted the depth and complexity of issues contributing to the health inequalities faced by this group which result in the greater rates of obesity (**Table 1**). These related to less finances available to put towards healthy food and activities, inequalities in access to healthy neighbourhoods, greater difficulties making healthy food choices in our unhealthy food environments, health issues and medications that may cause weight gain, difficulties with food textures or repetitive patterns of eating, difficulties cooking food, limited understanding of how food choices affect health, and the impact of comfort eating.

**Table 1:** Challenges to achieving and maintaining a healthy weight in people with intellectual disabilities, as identified by interview and focus group participants

| People with intellectual disabilities may: | Exemplar quote |
| --- | --- |
| Less finances available to put towards healthy food and activities | *“...food that’s cheap is the less healthy option. And if you’re on a short budget, and you haven’t got a lot of money, then it’s very, very tempting. (P03, social care team lead)*<br>*So I would say that’s a barrier: if people haven’t got access to do activities... now, instead of going swimming, bowling, walking to the local shopping centre, doing all different activities, he’s now sat in a chair most of the day... I say, I think that’s a barrier for people with learning disabilities. It depends on where they’re placed and what support they have. (P04, social care manager)* |
| Inequalities in access to healthy neighbourhoods | *“...the things that affect health inequalities, broadly speaking, I feel are exacerbated in the learning disability cohort. So they are probably more likely to live in areas of deprivation, probably less likely to have access to green spaces, less likely to have access to fruit and veg. Not always, but I think, statistically speaking, that’s what we tend to see on programmes...maybe 70% of our cohort in [city] are from the bottom two quintiles of deprivation. (P06, community weight management provider)* |
| greater difficulties making healthy food choices in our unhealthy food environments, for example being more influenced by food advertising | *"We have our more independent people, who go out on their own, and they go into town, and we have more problems with managing their healthy lifestyles...They're just so open to advertisements...their heads are turned by the pretty adverts." (P02, social care manager)* |
| Health issues and medications that may cause weight gain | *"The complexity of their physical, mental health...We have certainly seen much more coexisting mental health diagnosis as well alongside a learning disability...I think the two together adds an even higher level of complexity..." (P01, lead dietitian)*<br>*"We also have people on medication that makes them... We know risperidone makes the customers crave more carbs..." (P08, support worker)* |
| Difficulties with food textures or repetitive patterns of eating | *"I've supported lots of people who have put on weight as a result of a SALT [Speech And Language Therapy] diet because, all of a sudden, all of those healthy things, rice is a no-no, salad is a no-no. And staff are like, "Well, I don't know what to prepare," so it's easy to prepare foods that are now not nutritious anymore. (P16, social care manager, focus group)* |
| Difficulties cooking food | *"I think that's one of the challenges, is people live on their own and may not know how to cook, may not have ever been taught to cook. So, the push for them is takeaways and ready meals." (P12, healthcare commissioner)* |
| Limited understanding of how food choices affect health | *The understanding of good and bad food choices...It does make you feel better for a few moments...However, I have the understanding that, if I keep eating doughnuts and getting that feel-good feeling, I'm going to end up not being able to get up the stairs and play football with my grandchildren. So, it's around capacity. (P08, support worker)* |
| The impact of comfort eating | *"when a lot of people have been in care for a long time, they don't have much control over their life...And the only thing they do really have control over is what goes in their mouth, and food becomes a pleasure that they don't want to deny themselves with." (P02, social care manager)* |

##### Accessing and engaging with ‘mainstream’ weight management services

Participants also reflected on how people with intellectual disabilities who were experiencing challenges with their weight were able to access and engage with weight management interventions. Most participants viewed current mainstream weight management programmes as not accessible or suitable for people with intellectual disabilities. It was difficult to navigate the processes to even get a referral, particularly when it was via self-referral.

> *“…there are no easy reads, not a lot of explanations. Would I understand how to make a phone call and do a self-referral?” (P08, Support Worker)*

Getting a referral accepted by mainstream services was also challenging.

> *“It’s very difficult to get a mainstream service to accept somebody with complex learning disabilities…there’s a relatively low threshold for bouncing those referrals.” (P05, Manager, community learning disability healthcare team)*

There was a feeling that services weren’t commissioned or set up for people with more complex needs from the weight management provider perspective:

> *In an ideal world everybody would…have an offer based on their individual needs…But we don’t have those luxuries, and we are back to feeling constrained by our offer and having to serve the majority, which I am not saying is correct. (P01, lead dietitian)*
>
> *“In service specs, it’s “You need to make reasonable adjustments,” but no one defines what (those) are. (P06, Community weight management provider)*

However, a few felt that mainstream services could work well for some individuals with mental capacity who had adequate support to attend the services with them and help them put actions into practice.

> *We’ve had a guy…he had mitral valve regurgitation, so he needed to lose some weight…he made agreements with the dietitian, and then we helped him to follow them through…we had lots of very simple Easy Read, that we created with the dietitian…we went with him (P02, social care manager)*

Defining ‘success’ of weight management interventions

Overall participants felt that how success was defined in terms of weight management interventions needed to be re-thought in general, but particularly for people with added complexities such as intellectual disabilities. This included the importance of emphasizing non-scale victories and other health gains rather than just focusing on the scales.

> *I am increasingly frustrated with weight loss being the only outcome of whether something is effective because often a patient will not have lost any weight that I work with, but they have had loads of other really good outcomes that are not captured…Does mood improve, does self-worth improve? Maybe strength in terms of tying shoelaces…sometimes all those things happen and weight doesn’t change…(P01, lead dietitian)*

Participants described how it takes longer for people with intellectual disabilities to experience changes in weight and “requires additional support” over “a longer period of time” (P06, community weight management provider). The intersection of an intellectual disability with trauma and mental health issues was highlighted as adding to the complexity.

> *…many people we support have had trauma in their early years, so they’re dealing with these complicated psychological issues…we need to be mindful in those complex situations that there won’t be a quick fix. It takes time, perseverance, and ongoing. (P17, social care manager, focus group)*

Stopping weight gain and achieving weight maintenance was highlighted as a significant positive outcome, and some participants noted there was a tendency to focus on instant results.

> *“Our biggest achievement is that we’ve had weight maintained. That’s massive for us…And we have to take that as a win…Because people (support workers) get deflated, and they do give up too easily, if they don’t see instant results.” (P02, Social care manager)*

##### The role of the carer in supporting healthy weight

Overall, participants felt that it was within a carer’s role to support people with healthy weight in order to “support them holistically, to meet physical, emotional, social, medical needs” (P11, social care manager).

The power of positive role modelling was discussed – and many felt that carers did have a “huge responsibility to be good role models”. (P03, Social care team lead). One participant, however, felt that you couldn’t tell people to be a positive role model but at least they should put forward a positive message.

> *“You can’t say to someone, ‘You’ve got to be a positive role model…you need to be really healthy yourself,’…But it would be important that they still are a positive message to say to people, ‘It is important what you eat’” (P14, Social enterprise professional)*

Carers were described as holding ‘a lot of power’ in the relationship which also pertained to demonstrating and supporting healthy behaviours. This was particularly relevant if the carer was responsible for the shopping and preparing meals.

> *“If they’re the ones doing the shopping and preparing the meals, they have a lot of the power in that relationship” (P06, Community weight management provider)*

##### Challenges for carers in supporting healthy weight in adults with intellectual disabilities

Four key barriers to carers being able to support healthy weight in people with intellectual disabilities were highlighted. This included 1) wider financial and workforce pressures for carers 2) limited knowledge and skills related to healthy behaviours, 3) longstanding habits around using food as a reward for people with intellectual disabilities, and 4) perceived tension between supporting independence and choice and promoting healthy behaviours.

Wider financial and workforce pressures for carers

For paid carers, workforce pressures in the care sector were highlighted as a key issue such as lack of staff and poor pay, with healthy eating and physical activity not prioritised:

> *They’re short-staffed. They’re stretched…Weight management is the least of their problem. It’s, “Who’s coming in next?” I don’t think it’s up there on the list of problems to worry about. (P08, support worker)*

When people had a limited number of support hours, time spent on cooking was often an area that was reduced by carers in favour of convenience foods.

> *“…care visits are timed, so they only get a certain amount of time. And actually, if you can put a microwave meal in and then go do mzedication, do a bit of cleaning, help the person change then that’s usually what will happen. (P13, local authority commissioner)*
>
> *“…they’ve been encouraged to buy [ready meals] because that means less support time to cook a decent meal… (P17, social care manager, focus group)*.

Carer support was felt to be critical to help people with learning disabilities engage in weight management services, however, it was acknowledged that carer support wasn’t often adequate to make this work, particularly the lack of time and resources that carers had to convert information into Easy Read.

> *“It takes a long time to convert the information we need into Easy Read…It is massively time consuming. And when you’ve got 28 other people… it is resources, I think.” (P02, social care manager)*

Finances to put towards healthy food were also raised as an issue for people with intellectual disabilities and unpaid carers, with a lack of knowledge on how to prepare healthy meals on a budget.

> *“…it’s also a financial issue…Even if you teach them how to cook really nutritious, healthy meals, can they do that on a budget? And I don’t think that’s something many groups identify and do.” (P10, healthcare commissioner)*

Workforce turnover was also raised as an issue that could hinder healthy lifestyle changes being put into practice.

> *“It might be that clients came with a different support worker each week, and they might not be able to help the client to their full capacity because they were missing key information taught in previous weeks…it was very much, “Oh, well, I wasn’t here. I didn’t know anything about this. We haven’t had any paperwork come home with us, etc” (P07, public health professional)*.

Limited knowledge and skills related to healthy behaviours

Health literacy was described as a significant barrier for carers being able to support people with intellectual disabilities with healthy behaviours.

> *I think the biggest thing is they don’t know about nutrition themselves…If you don’t know anything about nutrition in your own personal life, then you’re not going to know anything about nutrition to care for someone else… (P25, social care team lead)*

Related to this was the “skills gap” related to cooking skills of carers who in many cases were responsible for meal preparation.

> *“…care staff get trained in first aid, food hygiene, communication, mental capacity, they’re never trained in cooking…If they’re a great cook you’re on to a winner, if they’re a bit rubbish you tend to go that way as well, so there’s a skills gap there. (P13, local authority commissioner)*

Carers feelings relating to their own journeys with weight and societal weight stigma were also felt to hinder discussions about healthy weight.

> *“I’ve worked alongside colleagues who are almost anti any discussion about healthy eating options or encouraging people because they’re defensive about their body shape and how they feel about that…I have witnessed that in the past where people were nervous to mention healthy options because somebody would take offence to that.” (P17, social care manager, focus group)*

Longstanding habits around using food as a reward for people with intellectual disabilities Participants described how there was a longstanding habit of using food as a reward for people with intellectual disabilities.

> *“Go on, have a piece of cake, it will make you feel better.” There is quite a lot of that messaging that goes out. Because even today, people are uncomfortable with disability*.
>
> *There’s a tendency to feel sorry, can’t do anything, but I can give you something nice… (P03, Social care team lead)*

Some participants felt there was a perception that because an individual had a disability, it didn’t matter if their weight was going up as their overall “quality of life was impaired anyway” (P03, social care team lead).

> *“It is okay that weight is going up because-Almost using the learning disability as a reason for making that okay, and does it matter” (P01, lead dietitian)*

Perceived tension between supporting independence and choice and promoting healthy behaviours

Carers talked about how if a person with an intellectual disability was deemed to have mental capacity to make decisions relating to healthy behaviours, there was very little they could do if the individual made unhealthy choices.

> *Yes, so it’s always harder when you’ve got somebody that has got capacity but doesn’t want to do something that would perhaps be in their best…He’s got that understanding, but then he’s saying, “I don’t want to lose weight,” in which case you always have to go with the person… (P04, social care manager)*

However, it was discussed that capacity was a complex topic particularly for long-term choices.

> *“I would argue that if somebody is consistently making unwise choices, regardless of whether they’ve got a disability or not, are they actually operating under capacity?…if you keep making that decision, and can’t stop, do you really have capacity?” (P03, social care team lead)*

A commissioner commented how the training emphasis was now so focused on the rights of the individual that basic health needs were forgotten:

> *“…a lot of training for formal paid carers, is around capacity…it promotes the rights, but the actual basic health needs are often probably forgotten. We’ve gotten so into people’s rights and independence and choice; they can choose to do this but are they necessarily well informed of the consequences of doing it?…weight loss is a long-term informed choice isn’t it? (P13, local authority commissioner)*

If a carer didn’t have the nutrition knowledge and cooking skills, then a real choice wasn’t necessarily available to people:

> *“…people are dependent on their support worker, often, to be the cook. If they’re a good cook, that’s brilliant, but if they are not a good cook…she doesn’t have a choice” (P14, social enterprise staff)*

It was felt that capacity was “a bit of a grey area” and could sometimes be used as a “fall-back…or get-out clause” (P14) for carers to not encourage people to make healthier choices.

From the perspective of a parent carer, a lot more positive guiding or persuasion was needed.

> *When I said, “I’d rather you didn’t allow him to buy anything to eat, he has had his packed lunch, he has had enough”, the retort I got was, “He’s over 18, he can make his own decisions, we can’t tell him what to do.” I said, “No, but you could perhaps persuade him that that wouldn’t be as good for him, and he has had his lunch…it’s how you present things to them but you have to be very good at persuasion as well.” (P23, parent carer)*

##### Carer training needs

Participants reported that a significant proportion of the UK care workforce have English as a second language and low literacy levels and stressed the importance of using simple language with visual prompts within any training. This, together with the context in which UK carers are working (see ‘Wider financial and workforce pressures’ above), highlighted the need for the e-learning to be chunked into small ‘bite sized’ sessions that could be engaged with during a break for example. Carer training needs related to the topic of healthy weight were categorised into three main areas forming the basis of e-learning sessions:

1. Understanding factors contributing to excess weight in people with intellectual disabilities.

Presenting simple facts and figures about the health inequalities in people with intellectual disabilities and how obesity impacts on health was seen to be important as it was felt that this was not well known.

> *“…giving them some facts and figures, how obesity impacts on people’s health… because some of them were jaw-dropping: ‘Really? They die 20 years younger?’“ (P12, healthcare commissioner)*

2. Understanding the basics of healthy eating and physical activity recommendations.

It was felt that many carers had a limited understanding of these basics, which meant this wasn’t passed onto the people being cared for.

> *“Staff don’t understand portion sizes…so they’re not passing that on to the customers…adults in the general population have a lack of understanding of food.” (P08, Support Worker)*

3. Understanding how to apply this knowledge when working with a person with an intellectual disability.

> *“…the tools of how you introduce that to somebody with a learning disability, who might not cope well with change” (P11, Manager, Social care)*

The national e-learning developed from this research is available at: https://www.e-lfh.org.uk/programmes/supporting-healthy-weight-in-an-adult-with-a-learning-disability/.

### Overarching concepts

This study highlights two overarching concepts central to understanding the challenges of supporting healthy weight in adults with intellectual disabilities: 1) Carers as central but under supported agents in health promotion, and 2) Structural and cultural barriers to healthy weight for people with intellectual disabilities. Carers are critical enablers of health promotion, yet they operate within systems that offer limited support, training, and resources. Their influence over food choices, daily routines, and access to services is substantial, but often constrained by workforce pressures, low health literacy, and tensions between promoting autonomy and guiding healthier behaviours. This together with socioeconomic disadvantage, inaccessible mainstream weight management services, and narrow definitions of success related to behaviour change contributes to the structural and cultural barriers to healthy weight for people with intellectual disabilities.

## Discussion

People with intellectual disabilities face multiple health inequalities that increase their risk of obesity. These relate to wider determinants of health, co-existing conditions, and reliance on carers to support healthy lifestyle choices. Weight management services are often poorly accessible requiring additional time and resources to engage individuals and carers effectively. Carers play a critical role in supporting healthy behaviours in people with intellectual disabilities; however, they face many barriers in enabling this. These include financial and workforce pressures, limited knowledge of obesity, nutrition and physical activity, weight stigma, habits around using food as a reward, variability in cooking skills, and a perceived tension that promoting healthy choices is incompatible with the rights and independence agenda. This project sought to investigate these issues in depth and address identified education needs through development of a three-session, freely available, national <u>e-learning</u> package for carers on how to support healthy weight in people with intellectual disabilities.

Our finding on perceived poor accessibility of mainstream weight management services for people with intellectual disabilities aligns with a recent systematic review and meta-analysis by Rana *et al* (2024), which examined lifestyle modification interventions targeting health risk behaviours in adults with intellectual disabilities (8). The review found physical activity and dietary interventions had limited effectiveness in changing behaviours or achieving weight change in this group. The authors called for population specific strategies, measures and evaluation frameworks, co-produced with people with lived experience (8). Currently, bespoke weight management services for people with intellectual disabilities are not routinely commissioned in the UK.

Croot *et al* (2018) undertook a qualitative study with people with intellectual disabilities, their carers, and staff from a commercial weight loss programme, to identify adjustments to improve accessibility for people with intellectual disabilities (13). They found carer engagement was critical to facilitating behaviour change even when reasonable adjustments, such as simplified information and staff training, were made. Rana *et al* similarly highlighted carers’ essential roles in facilitating recruitment, communication, and implementation of lifestyle interventions, recommending that future studies designing lifestyle interventions target both people with intellectual disabilities <u>and</u> carers and document the level of carer support received when evaluating the intervention (8). Taken together, this literature highlights a role for bespoke weight management programmes for people with intellectual disabilities designed in conjunction with people with intellectual disabilities and carers.

Knowledge gaps around obesity, healthy eating and physical activity and the need for training have been highlighted in previous UK studies by Croot *et al* (2018) (qualitative interviews with carers) and Melville *et al* (2009) who undertook a cross-sectional interview-administered questionnaire of 63 paid carers (13, 23). Similar findings have been reported in Norway and Sweden, based on qualitative methods with carers (24, 25). These studies found that nutrition education and meal-oriented support was not prioritised, and carers lacked time and resources to involve residents in cooking, with only the most ‘necessary’ tasks prioritised. Both studies highlighted that structural and organisational changes may be needed to prioritise healthy diet for people with intellectual disabilities.

The tension between supporting independence and choice and promoting healthy behaviours has been reported in several studies (13, 23, 24, 26, 27). Smyth & Bell argued the focus on promoting choice could inadvertently lead to encouraging unhealthy choices and reasoned that if an individual is not taught about a healthy alternative that is available to them, they lack real choice (26). They highlight the importance of exploring whether someone truly understands the consequences of a choice and is therefore able to make a true choice. This topic around mental capacity and choice are included as a key area in session 3 of our e-learning. Other studies have reported that food choices were often limited to a few options pre-selected by carers or service providers, influenced by staffing resources, and carer knowledge, skills and preferences (13, 24, 25). Professor Chris Hatton argues that supporting independence and choice is not incompatible with promoting healthy behaviours and contends that the limited range of unhealthy options offered inevitably results in ill health being the only choice (27). Despite being reported in the literature over the past two decades with repeated calls for action, this issue remains largely unaddressed underscoring an urgent need for change (13, 23, 27).

A strength of this research was the in-depth qualitative approach which enabled exploration of the complex barriers and training needs in the care sector, from a multi-disciplinary social care, healthcare, public health, commissioning and voluntary sector perspective. The iterative design allowed refinement of training content and incorporation of real-life examples to ensure the training was relevant and grounded in current issues. Partnering with a learning disability charity allowed us to bring to life the themes of the research through lived experience videos. This research was commissioned by NHS England and was therefore limited to participants in England. Although we anticipate that many findings will be transferable to other countries, adaptation of the training may be needed for use outside of England.

Given the disproportionate health burdens and premature death linked to obesity in people with intellectual disabilities, reducing health inequalities related to obesity should be a key research and policy priority. Achieving this requires a range of actions underpinned by research (9). Carer training is one element addressed in this research. The training developed is introductory and should be built on to address topics in more detail, in particular, balancing independence and choice with promoting healthy behaviours and duty of care. Weight management services should be co-designed with people with intellectual disabilities and carers to improve carer engagement and likelihood of success. Future research should also consider intersectionality and overlapping health inequalities related to obesity.

## Conclusions

In conclusion, our findings reinforce the central role that carers play in supporting healthy behaviours among people with intellectual disabilities, while highlighting the significant and multifaceted barriers they face in doing so. Addressing these challenges, through targeted training, structural support, and co-designed interventions, is essential to improving accessibility and effectiveness of weight management support for people with intellectual disabilities. Our e-learning programme offers a foundation for carer training in this area, but further development and integration into wider systems is needed. Reducing obesity-related health inequalities in this population demands a sustained, inclusive approach that empowers carers and respects the rights and choices of people with intellectual disabilities.

## Data Availability

Anonymised interview and focus group transcripts from this research will be deposited in the University of Bristol Research Data Repository. These anonymised data can only be accessed through approval of a data access committee.

## Notes

### Competing Interest Statement

KC has previously undertaken research consultancy for Oviva, Manual and Oxford Medical Products through University of Bristol research consultancy agreements ? these are unrelated to the research reported in this manuscript. CA is a trustee for the Caroline Walker Trust. The Caroline Walker Trust allowed the use of selected food photos to be used within the e-learning developed from this research.

### Funding Statement

This research was funded by a grant from Health Education England. The funders had no role in study design, data collection and analysis, decision to publish, or preparation of the manuscript.

